# The effect of physical activity on brain structure and cognitive function in the population-based cohort of LIFE-Adult-Study

**DOI:** 10.1101/2025.10.23.25338642

**Authors:** Polona Kalc, Rober Dahnke, Christian Sander, Frauke Beyer, Andrea Zülke, Steffi G. Riedel-Heller, Veronica Witte, Christian Gaser

**Author notes:** Corresponding author: Polona Kalc, Address: Am Klinikum 1, 07747 Jena. Co-senior authors (Veronica Witte, Christian Gaser).

## Abstract

Physical activity is believed to positively influence brain health and cognition and is considered a modifiable lifestyle factor that may protect against cognitive decline and neurodegeneration. In this observational study, we investigated the cross-sectional and longitudinal effects of self-reported total and moderate-to-vigorous physical activity on cognitive scores on the Trail Making Test (TMT-A and TMT-B), hippocampal volume, and BrainAGE, in a large population-based cohort from the LIFE-Adult Study (*n* = 2576). Furthermore, we examined the effect of objectively measured physical activity on brain structure in a subgroup with available accelerometry data (*n* = 227). Multiple linear regression analyses did not show any positive effects of self-reported or objectively measured physical activity on hippocampal volume or processing speed and executive function. Longitudinal path analyses suggested a potential for reverse causation, where a higher BrainAGE at baseline was associated with lower physical capacity at follow-up. Additionally, we observed an age-related bias in the self-reporting of physical activity, indicating that older individuals tend to overestimate their level of activity. Future interventions targeting middle-aged adults may be necessary to raise awareness of potential misperception and encourage increased physical activity.

## Introduction

Physical activity, defined as any behaviour involving bodily movement that results in energy expenditure and increased physical fitness [1, 2], and its subcategory of physical exercise, which is movement performed in a structured and repeated manner in order to improve physical fitness [1], have emerged as promising modifiable lifestyle factors that support healthy development and ageing [3, 4]. In the context of neuroscience and brain health, physical activity and exercise have been identified as key lifestyle factors that influence brain plasticity [5, 6], with accumulating evidence suggesting potential for delaying cognitive decline and reducing the risk of dementia[7–9].

A large body of observational and experimental studies reports beneficial effects of physical activity on brain structure and function across the lifespan [10, 11]. Evidence from animal models has identified several mechanisms through which physical activity may support brain health and cognitive function, including the increased production of neurotrophic factors such as brain-derived neurotrophic factor, enhanced cellular signalling promoting neural plasticity, and neurogenesis [12, 13]. Neuroimaging research has demonstrated the associations of physical activity with white matter integrity [14, 15] and grey matter volume, predominantly in the (left) hippocampus [16, 17], but also in cerebellum [18], frontal, and parietal regions [19]. Several studies using machine learning algorithms to derive markers of brain ageing have also linked physical activity to younger-appearing brains [20–22]. Cognitive function has similarly been found to benefit from physical activity, particularly in domains such as executive function, memory, and processing speed [23, 3, 24].

Nevertheless, a recent large-scale population-based study using the UK Biobank cohort reported negligible effects of physical activity on cognitive function, potentially due to sampling bias (volunteer-based sample), younger sample age, or reverse causality [25]. Similarly, no significant relationship was observed between physical activity and the neuroimaging-derived biomarker of brain ageing, the so-called “brain age gap”, within the same cohort [26]. In light of these contradictory findings, the present study aimed to estimate the cross-sectional and longitudinal effects of physical activity on brain structure and cognitive function in the population-based LIFE-Adult cohort of individuals from Leipzig, Germany [27, 28]. Based on a large volume of research, we hypothesised that there would be a positive effect of physical activity (both self-reported and assessed by accelerometer) on brain structure and cognitive function. BrainAGE (Brain Age Gap Estimate), a machine learning derived marker of structural brain aging, and hippocampal volume were used as brain structural outcomes, and processing speed and executive function (derived from Trail Making Test A & B, respectively) as cognitive outcomes. Additional analyses were conducted to address the potential age-bias effect and a quasi-cross-lagged panel model was estimated to investigate the possibility of reverse causality.

## Methods

### Participants

The participants in this study were part of the LIFE-Adult Study, a large population-based cohort study, whose baseline assessment was conducted from August 2011 to November 2014 by the Leipzig Research Centre for Civilization Diseases (LIFE) [28]. The LIFE-Adult Study was approved by the ethical board of the Medical Faculty of the University of Leipzig. All the participants provided their written informed consent and received a small remuneration for their participation.

Ten thousand randomly selected participants from Leipzig, Germany, underwent an extensive data collection at baseline, consisting of physical examinations, anthropometric data collection, interviews on medical and familiar history, self-report sociodemographic and lifestyle questionnaires, as well as cognitive testing [28]. Subsequently, a subset of participants wore an accelerometer to assess physical activity and sleep patterns. Structural and functional neuroimaging data were acquired for a large proportion of participants at a separate visit several weeks after the baseline visit of the study.

The follow-up data of 1799 participants were collected from June 2018 to August 2021 with as much overlap between the measures from the first time point as possible [27]. Accelerometer data were not collected at the follow-up.

In the present study, the analyses were conducted on a base-line sample of 2576 participants (age range: 19-82, Mdn = 64.58 years, IQR = 22.41 years, 47% women). Two separate longitudinal path analyses were performed on two (overlapping) smaller subsamples for which data on physical activity was available at both time points (sample with cognitive scores: *n* = 342, Mdn = 63.57 years, IQR = 27.38 years, 40.1% women; and sample with structural BrainAGE: *n* = 253, Mdn = 62.88 years, IQR = 30.42 years, 39.5% women).

**Figure 1.**
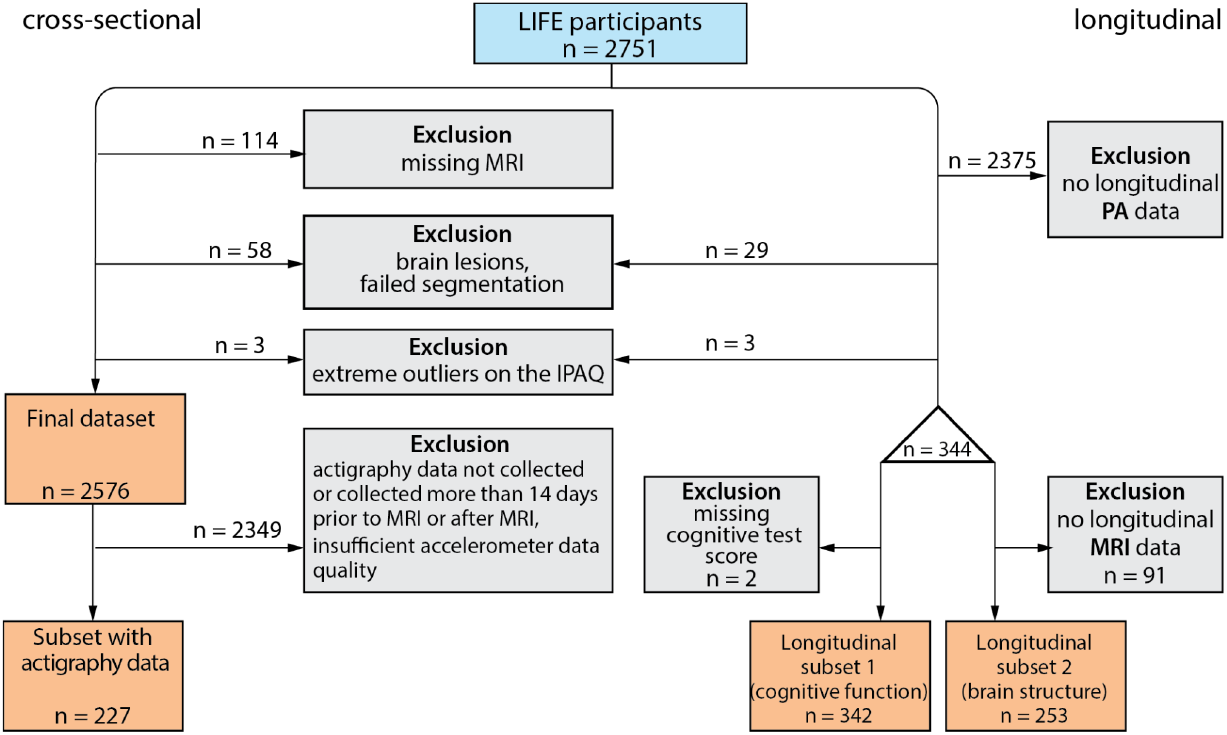
Flowchart of data selection for cross-sectional analyses (left) and longitudinal analyses (right).

### Measures

#### Exposure: Physical Activity

Physical activity at baseline was measured using a short version of the International Physical Activity Questionnaire (IPAQ-SH, [29]), a self-report measure of physical activity over the previous 7 days. Participants recorded how many days in the previous week they had spent walking or performing moderate or vigorous physical activities, as well as how many hours and minutes they had spent at each respective intensity level on each day. The evaluation was carried out based on the published evaluation guidelines (https://sites.google.com/view/ipaq/score), calculating the sum of the three reported activity areas: walking, moderate physical activity, and vigorous physical activity. The resulting measure is expressed in metabolic equivalent of task (MET) minutes per week [30]. In this study, we examined the effects of both total physical activity (the sum of all reported physical activities) and moderate to vigorous physical activity (the sum of moderate and vigorous physical activities), as previous studies have shown that moderate-to-vigorous, rather than total, PA drives changes in brain structure and/or function [31, 21, 32].

A subset of participants who were willing to participate in the sleep and activity part of the LIFE-Adult study also wore a 2D-axial accelerometer SenseWear Pro 3 (Bodymedia, Inc., Pittsburgh, Pennsylvania). Participants were asked to wear the accelerometer for seven consecutive days. Only the data from participants who had worn the accelerometer for at least four weekdays and one weekend day, and at most 14 days prior to the scanning session, were used in the analysis. The raw actigraphy data were processed using the SenseWear Professional software package version 6.1. The minute-by-minute data was subsequently exported to a Microsoft Excel template. The Excel template was used to customise the analysis windows according to the specific day-night-cycles of each participant (the standard SenseWear software uses a fixed time-window for activity analyses, e.g. 12 am to 11:59 pm). Based on the bedtime information provided in a sleep diary kept during the actigraphy recording, the respective night sleep intervals were identified manually. For the subsequent analyses, only daytime intervals with a minimum duration of 20 hours and an off-wrist duration of less than 15% of the daytime interval were used. Activity parameters were then recalculated for the daytime interval (defined as the period between two consecutive night sleep intervals). We used a measure of energy expenditure in MET-minutes that were individually averaged across the measuring days for the participants.

During the follow-up, physical exercise capacity rather than physical activity was measured using the Veteran Specific Activity Questionnaire (VSAQ, [33]). This questionnaire consists of a list of 13 activities arranged in order of increasing physical demand (e.g., from “Eating”, “Taking a shower” to “Any competitive activity”). Participants had to rate which activity would result in notable fatigue and heavy breathing. Their exercise capacity in MET was extrapolated based on the self-reported loading and age [33, 34].

#### Outcome: Brain structure

Brain structure was assessed from T1-weighted MRI scans, acquired on one 3T Siemens Verio scanner with a 32-channel coil. An MP-RAGE sequence with 1 mm isotropic voxels, 176 slices, TR=2300 ms, TE = 2.98 ms, and inversion time (TI) = 900 ms was used. The images were processed with the CAT12.9 [35] of SMP12 in Matlab 2024b. Two structural outcomes were devised: BrainAGE and hippocampal volume. The quality of the images was automatically tested using the CAT12 structural image quality rating (SIQR, [36]), with a cut-off threshold of 2.4, and by visual inspection, using the ‘Check Sample Homogeneity Tool’ in CAT12.

#### Brain age gap estimation

The Brain Age Gap Estimate (BrainAGE) represents a difference between a person’s chronological age and the age estimated by machine learning based on their brain scan. A positive or negative BrainAGE indicates that the brain appears older or younger than expected for the person’s age, respectively [37, 38]. The BrainAGE machine learning model (Gaussian process regression) previously described in Kalc et al. [39] was trained on a subset of scans from a large sample of healthy participants from various openly available neuroimaging datasets (*n* = 2446, M_Age_ = 57.69 ± 14.88 years, 58% women). It was then applied to baseline and follow-up brain images from the LIFE-Adult sample. As the brain age gap is always overestimated for younger subjects and underestimated for older ones, an age-bias correction was applied as described in Cole [26], resulting in a mean absolute error of 4.68 years and a correlation to age of 0.91. The age-bias correction is used to remove the dependency of the predicted brain age on age [40]. Therefore, we did not further control for the age in the regression analyses using the BrainAGE as the outcome. A detailed process of training and testing the algorithm is available in the Supplement.

#### Hippocampal volume

Hippocampal volume was extracted from the segmented grey matter regions using anatomical definitions provided by the Neuromorphometric atlas (Neuromorphometrics, Inc.). Specifically, the left and right hippocampal regions were identified and normalised by the total intracranial volume based on the analysis of covariance approach as described in Raz et al. [41]. The adjusted volumes were averaged into a single bilateral hippocampal volume measure.

#### Outcome: Cognitive function

Cognitive functioning of participants was assessed by a set of neuropsychological tests [28]. In this study, the time-to-complete scores from the Trail Making Test A and B (TMT, [42]) were available for both time points for the majority of the participants, and were therefore used as cognitive outcome measures. In TMT-A, participants had to connect the numbers on the paper in the correct order as quickly as possible. In part B, they had to connect numbers and letters of the alphabet, correctly alternating between both. The maximum time allowed for completion was 150 seconds and 300 seconds for part A and B, respectively. The time-to-complete scores (without accounting for errors) were used as a measure of processing speed and executive function of the participants (scores on TMT-A and TMT-B, respectively).

#### Covariates

Our covariate selection was based on the conceptual model of Campbell et al. [43], including sociodemographic factors and adult health behaviours. However, several possible covariates, such as factors that occurred prior to the birth or earlier in life, were not available in our dataset, therefore we opted for a smaller number. A minimal adjustment set of covariates was tested in the *dagitty* toolbox [44] (Figure S1 in the Supplement). Linear (and quadratic) polynomial terms of age, along with tobacco exposure (i.e., present and past smoker vs. non-smoker), alcohol consumption (g/day) and socioeconomic status (dummy coded) were included as covariates to account for confounding. Multicollinearity of the covariates was assessed using the formula for variance inflation factor: VIF = 1 / (1 - R^2^). The VIFs indicated no signs of multicollinearity, with all values being ≤ 1.757.

### Statistical analysis

Statistical analyses were performed in R 4.4.3 [45], using RStudio version 2024.12.1+563. The cross-sectional multiple linear regression models were analysed with *lavaan* package version 0.6-19 [46], using a robust maximum likelihood estimation (MLR). Standard errors were calculated using a robust (Huber-White sandwich) estimator to better account for violations of normality. Missing data were handled by full information maximum likelihood (FIML) with the assumption that data was missing at random. We estimated the effect of self-reported total as well as moderate to vigorous physical activity as MET hours/day on two structural brain outcomes (i.e., BrainAGE and hippocampal volume) and two cognitive outcomes (i.e., attention and cognitive flexibility). As cognitive tests were performed before the accelerometer data collection, cognition was not included as an outcome in the analysis with accelerometer-derived physical activity measure.

Two path analyses with a quasi cross-lagged panel design were estimated for the subjects with available longitudinal physical capacity, BrainAGE, and cognitive scores to analyse the potential reverse causality.

## Results

Sample characteristics for a cross-sectional baseline sample and its subsample with available actigraphy data, as well as for the two longitudinal subsamples with available cognitive scores (subsample 1) and structural MRI (subsample 2) are available in Table 1.

**Table 1.** Sample characteristics.

|  | Total baseline sample<br>(n = 2576) | Actigraphy subsample<br>(n = 227) | Longitudinal subsample 1<br>(n = 342) | Longitudinal subsample 2<br>(n = 253) |
| --- | --- | --- | --- | --- |
| **Demographics** |  |  |  |  |
| Age [years]<br>Mdn (Q1, Q3) | 64.47<br>(48.06, 70.60) | 65.93<br>(50.64, 71.32) | 63.57<br>(42.47, 69.86) | 62.88<br>(38.79, 69.21) |
| Sex (% women) | 1204 (47%) | 110 (48%) | 137 (40%) | 100 (39.5%) |
| Missing (%) | 4 (0.16%) | 0 (0%) | 0 (0%) | 0 (0%) |
| **Physical activity** |  |  |  |  |
| Total PA [MET min/week]<br>Mdn (Q1, Q3) | 4056<br>(2133, 6750) | 4074<br>(2430, 6510) | 4158.75<br>(2106, 6684) | 4293<br>(2106, 7038) |
| Missing (%) | 405 (15.7%) | 27 (11.9%) | 0 (0%) | 0 (0%) |
| MVPA [MET min/week]<br>Mdn (Q1, Q3) | 2400<br>(960, 4800) | 2640<br>(1200, 4320) | 2640<br>(960, 5040) | 2880<br>(760, 5280) |
| Missing (%) | 405 (15.7%) | 27 (11.9%) | 0 (0%) | 0 (0%) |
| **Lifestyle** |  |  |  |  |
| Smoking status (% smokers) | 1101 (43%) | 83 (37%) | 158 (46%) | 119 (47%) |
| Missing (%) | 105 (4.08%) | 5 (2.20%) | 2 (0.6%) | 2 (0.8%) |
| Alcohol consumption [g/day]<br>Mdn (Q1, Q3) | 4.75<br>(1.12, 16.14) | 5.52<br>(1.15, 18.32) | 5.70<br>(1.19, 16.87) | 6.17<br>(1.41, 16.66) |
| Missing (%) | 173 (6.72%) | 13 (5.73%) | 15 (4.4%) | 12 (4.7%) |
| **Socioeconomic status** |  |  |  |  |
| Low | 475 (18.4%) | 29 (12.8%) | 49 (14.3%) | 38 (15.0%) |
| Middle | 1530 (59.4%) | 144 (63.4%) | 207 (60.5%) | 149 (58.9%) |
| High | 548 (21.3%) | 52 (22.9%) | 85 (24.9%) | 65 (25.7%) |
| Missing (%) | 23 (0.89%) | 2 (0.9%) | 1 (0.29%) | 1 (0.40%) |
| **Cognitive function** |  |  |  |  |
| TMT-A time [s]<br>Mdn (Q1, Q3) | 34<br>(27, 44) | 33<br>(26, 43) | 33<br>(25, 44) | 32<br>(24, 42.25) |
| Missing | 18 (0.7%) | 1 (0.44%) | 0 (0%) | 1 (0.40%) |
| TMT-B time [s]<br>Mdn (Q1, Q3) | 78<br>(59, 103) | 76<br>(58, 96) | 76<br>(59, 96.75) | 71.5<br>(56, 94) |
| Missing | 29 (1.13%) | 1 (0.44%) | 0 (0%) | 1 (0.40%) |
| MMSE<br>Mdn (Q1, Q3) | 29<br>(28, 30) | 29<br>(28, 30) | 29<br>(28,30) | 29<br>(28, 30) |
| Missing | 1085 (42.1%) | 88 (38.8%) | 125 (36.5%) | 101 (39.9%) |
| **Brain structure** |  |  |  |  |
| Average hippocampal volume [ml]<br>M (SD) | 3.10<br>(0.25) | 3.09<br>(0.24) | 3.12<br>(0.22) | 3.13<br>(0.22) |
| Missing | 0 (0%) | 0 (0%) | 10 (2.9%) | 0 (0%) |
| BrainAGE<br>M (SD) | -1.31<br>(6.26) | -1.63<br>(6.43) | -1.29<br>(6.27) | -1.52<br>(6.17) |
| Missing | 0 (0%) | 0 (0%) | 10 (2.9%) | 0 (0%) |

The following section presents the effects of physical activity on brain structural and cognitive outcomes in the cross-sectional analyses. We show the standardized estimates from unadjusted and adjusted models reflecting the effect of self-reported total PA on brain structure and cognitive outcome in Table 2. The effects of objectively measured PA on brain structure can be seen in Table 3. We present the effects of self-reported amount of MVPA on brain structure and cognition in Table 4.

**Table 2.**
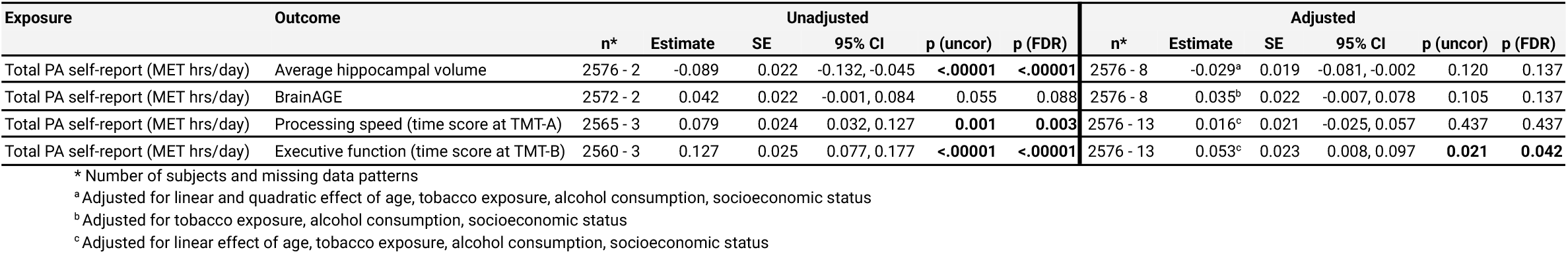
Unadjusted and adjusted cross-sectional parameter estimates of self-reported total physical activity on brain structure and cognition.

**Table 3.**
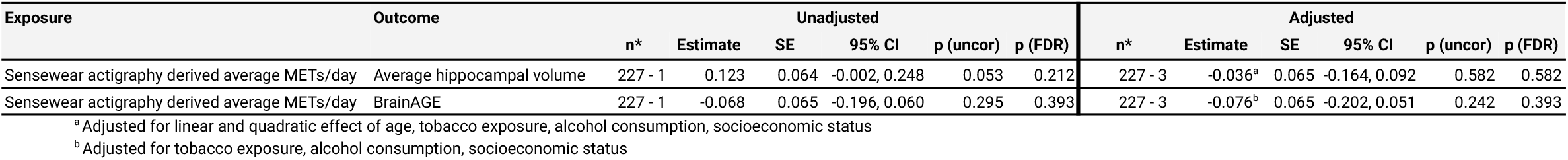
Unadjusted and adjusted cross-sectional parameter estimates of accelerometer-derived average MET/day on brain structure.

**Table 4.** Unadjusted and adjusted cross-sectional parameter estimates of self-reported moderate to vigorous physical activity (MVPA) on brain structure and cognition.

| Exposure | Outcome | Unadjusted |  |  |  |  |  |  | Adjusted |  |  |  |  |  |
| --- | --- | --- | --- | --- | --- | --- | --- | --- | --- | --- | --- | --- | --- | --- |
|  |  | n* | Estimate | SE | 95% CI | p (uncor) | p (FDR) |  | n* | Estimate | SE | 95% CI | p (uncor) | p (FDR) |
| MVPA self-report (MET hrs/day) | Average hippocampal volume | 2576 - 2 | -0.072 | 0.022 | -0.115, -0.028 | **0.001** | **0.004** |  | 2576 - 8 | -0.022<sup>a</sup> | 0.019 | -0.058, -0.015 | 0.245 | 0.280 |
| MVPA self-report (MET hrs/day) | BrainAGE | 2572 - 2 | 0.044 | 0.022 | 0.001, 0.087 | **0.044** | 0.072 |  | 2576 - 8 | 0.038<sup>b</sup> | 0.022 | -0.005, 0.081 | 0.080 | 0.107 |
| MVPA self-report (MET hrs/day) | Processing speed (time score at TMT-A) | 2565 - 3 | 0.058 | 0.025 | 0.009, 0.107 | **0.02** | 0.053 |  | 2576 - 13 | 0.011<sup>c</sup> | 0.021 | -0.030, 0.052 | 0.598 | 0.598 |
| MVPA self-report (MET hrs/day) | Executive function (time score at TMT-B) | 2560 - 3 | 0.101 | 0.025 | 0.051, 0.150 | **<.00001** | **<.00001** |  | 2576 - 13 | 0.045<sup>c</sup> | 0.022 | 0.001, 0.089 | **0.045** | 0.072 |

<sup>a</sup> Adjusted for linear and quadratic effect of age, tobacco exposure, alcohol consumption, socioeconomic status<sup>b</sup> Adjusted for tobacco exposure, alcohol consumption, socioeconomic status<sup>c</sup> Adjusted for linear effect of age, tobacco exposure, alcohol consumption, socioeconomic status

The results indicate no statistically significant effects of self-reported total PA on brain structure (β = -0.029, *p* = .137 and β = 0.035, *p* = .137 for hippocampal volume and BrainAGE, respectively). There is a statistically significant effect of total self-reported PA on cognitive function, indicating that higher levels of PA lead to higher time scores on TMT-B (β = 0.053, *p* = .042). Similar results can be observed for MVPA, however, the results do not survive the correction for multiple comparisons (β = 0.045, *p* = .072).

The analysis of objectively measured PA indicated no statistically significant effects on brain structure (β = -0.036, *p* = .582 and β = -0.076, *p* = .393 for hippocampal volume and BrainAGE, respectively).

The longitudinal effects of physical activity on cognition (executive function measured by TMT-B) and brain structural ageing (operationalised by BrainAGE) were estimated using path analysis. Self-reported total PA at baseline was used as a predictor of cognitive score or BrainAGE, and physical capacity at follow-up. The model testing the effects of PA on cognition showed no statistically significant effects of either PA at baseline on cognition at follow-up (β = 0.19, *p* > .05), nor of cognition at baseline on follow-up physical capacity (β = -.09, *p* > .05). The model demonstrated acceptable fit to the data according to robust fit indices: χ^2^ (1) = 2.21, *p* = .138, CFI = 0.995, TLI = 0.955, RMSEA [90% CI] = 0.077 [0.000, 0.22], SRMR = 0.026. In the model testing the effect of PA on brain structural ageing, we observed the effect of brain structural ageing at baseline on physical capacity at follow-up (β = -.11, *p* < .05), but not the effect of PA at baseline on brain structural changes at follow-up (β = -.01, *p* > .05). The robust fit indices of the model also suggested an acceptable fit (χ^2^ (1) = 1.08, *p* = .300, CFI = 1.000, TLI = 0.998, RMSEA [90% CI] = 0.019 [0.000, 0.187], SRMR = 0.025). The path diagrams are shown in Figure 2.

**Figure 2.**
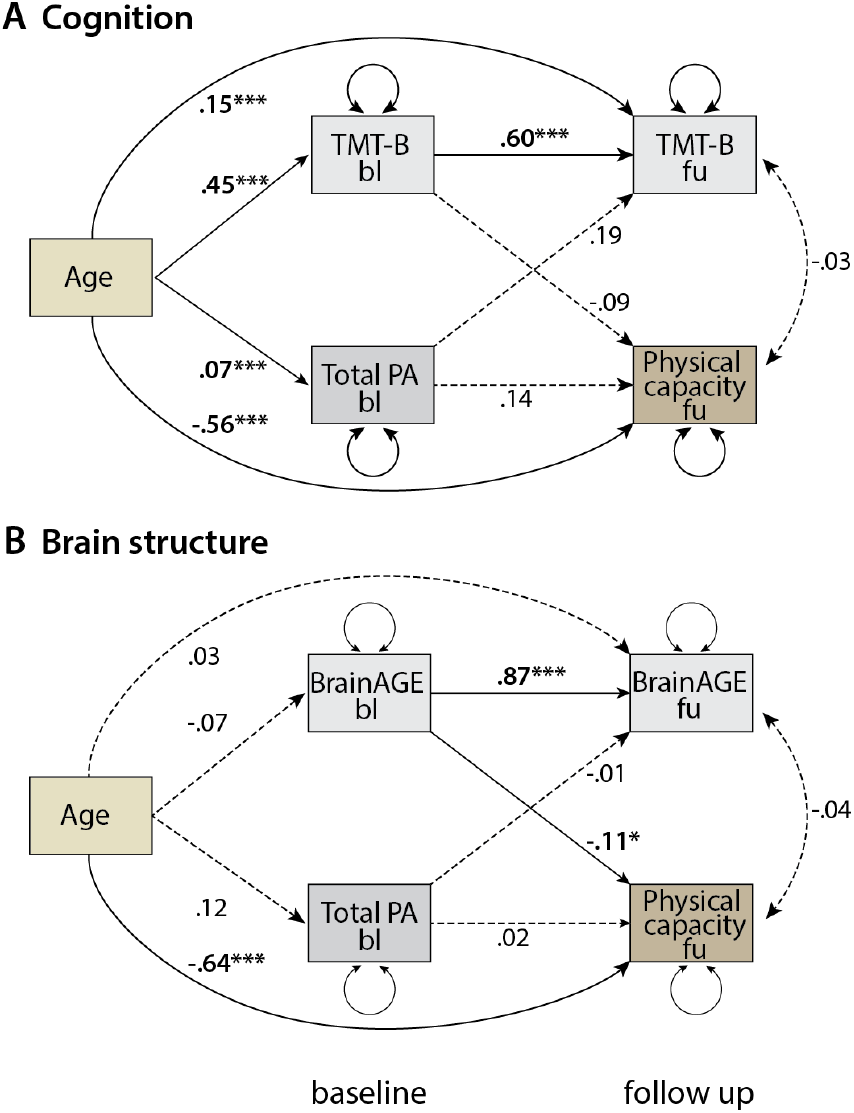
Graphical representation of two path analyses of the relationship between physical activity and A) cognitive function and B) BrainAGE at baseline, and physical capacity and cognitive function or BrainAGE at follow-up. Age at baseline was used as covariate at both time points. No statistically significant effects of either PA at baseline on cognition at follow-up, nor of cognition at baseline on follow-up physical capacity can be observed. Model B shows a negative effect of structural brain ageing on physical capacity at follow-up, but no effect of PA at baseline on brain structural ageing pattern at follow-up. (Note. * *p* < .05; \*\**p* < .01; *** *p* < .001)

### Sensitivity analyses

Given prior reports of lateralized effects of physical activity on left hippocampal volume [17], we conducted additional analyses focusing on this region only. However, no such effect was observed in our sample (β = -0.026, *p* = .269 and β = -0.046, *p* = .487 for self-reported total PA and objectively measured PA, respectively). The results are available in the Supplementary table (Table S1).

Furthermore, due to the unexpected negative association between the hippocampal volume and self-reported PA, we examined the potential self-report bias. We compared the correlation coefficients between self-reported total and accelerometer-derived PA in the samples of subjects below and above the age of 60 using the package *diffcor* [47]. The age of 60 was chosen based on the characteristics of the sample, and to roughly divide between the working and retired population. There were significant differences in the correlation coefficients between the two groups (*r*_<60_ = .52, *r*_>60_ = .29, mean difference = 0.28, 95% CI [.002, .525]), showing systematic bias in self-reporting measures of PA in younger vs. older participants. We therefore conducted the analyses for the two age groups separately. The results are available in supplementary Tables S2-S7. Self-reported MVPA showed no effect on brain structure and cognition in either age group, and total self-reported PA was positively associated with higher BrainAGE (β = 0.065, *p* = .05) and lower executive function (β = 0.083, *p* = .048) only in the older subsample. However, when examining objectively measured PA, we observed a moderate effect of PA on the younger-appearing brain in the subset of adults aged between 30 and 60 years (n = 70) (β = -0.343, *p* = .002), but we could not see the same effect in individuals over 60 years old (β = -0.006, *p* = .954) (see Tables S4 and S7).

Furthermore, we stratified the analyses based on biological sex. The results for men showed a small, statistically non-significant negative association of total self-reported PA with hippocampal volume (β = -0.047, *p* = .082). Similar results were observed for objectively measured PA in men (β = -0.072, *p* = .736 and β = 0.03, *p* = .760 for hippocampal volume and BrainAGE, respectively), with inverse signs observed for women (β = 0.071, *p* = .462 and β = -0.148, *p* = .115 for hippocampal volume and BrainAGE, respectively). Overall, the effects were small and none survived the correction for multiple comparisons. Full results are available in the Supplementary material in tables S8-S13.

## Discussion

A growing body of research shows the positive influence of physical activity on brain health and cognition [10, 11, 31]. In the present study, we investigated the effects of physical activity on brain structure and cognitive outcomes in a large, population-based cohort from the LIFE-Adult Study. We expected self-reported total and moderate to vigorous physical activity at baseline to have a positive effect on hippocampal volume and on making the brain appear younger (operationalised by smaller BrainAGE), as well as on better cognitive performance on the Trail Making Test. However, the results of multiple regression analyses did not support the hypothesised effect. Instead, the estimates showed the opposite direction (higher self-reported physical activity was negatively associated with hippocampal volume and positively with BrainAGE), although these effects were not statistically significant. There was a statistically significant negative effect of total self-reported physical activity on executive function, as measured by the TMT-B, showing an unexpected relationship between the exposure and outcome.

Self-reported measures of physical activity can be subject to reporting bias [48, 49]. A recent study examining sex-specific effects of self-reported physical activity on brain grey matter volume proposed that self-report measures of physical activity may not be sufficiently sensitive to investigate associations with brain health [50]. In line with this research, we found no effects of self-reported physical activity on hippocampal volume or structural brain ageing. Furthermore, we did not find any effects in sex-stratified sensitivity analyses. However, we observed an age-related difference in the association between the self-reported physical activity measure and the objectively measured physical activity, suggesting a measurement error in the predictor, which probably affected our results. This age-dependent reporting bias was previously demonstrated by other studies, where higher age was associated with overreporting activity levels [51-54]. Overreporting could stem from worsening recall, socially desirable responses, and the subjective nature of self-report questionnaires, which also depend on a person’s physical fitness [51]. Future (observational) studies would greatly benefit from including both accelerometer and self-reported measures of physical activity.

To overcome the limitations of self-reported measures, we examined the effect of objectively measured physical activity by accelerometer on brain structure for a subset of participants. We observed no statistically significant effect of accelerometry-derived physical activity on either hippocampal volume (β **=** -0.04, *p* = .582) or BrainAGE (β = -0.076, *p* = .242). This is in contrast to some large-scale accelerometer studies, showing positive association of physical activity on brain structure [55, 56, 21]. However, the reported effects in those studies were small, so it is likely that our subsample lacked the statistical power to detect them. Furthermore, the subsample with accelerometer data in this study exhibited rather low activity levels [30], which may have fallen below the threshold of intensity and/or duration that the literature identifies as necessary to observe beneficial effects on brain health [57, 58]. Our sensitivity analyses have shown that in a subset of participants younger than 60 years, higher accelerometer-derived activity was associated with younger-appearing brains (lower BrainAGE), although these participants were on average more active than the older group and we could not see similar effects in a larger older group.

Contrary to a large body of research showing the positive influence of physical activity on brain structure and function, our results suggest no consistent effects in a large cohort of citizens from Leipzig, Germany. However, although our dataset was predominantly older, it had a larger age range than those of other studies, which may have affected our results. To date, only a small number of studies have reported no effects of physical activity on brain health or cognitive function (e.g., [50, 59–61]). Some of those studies were conducted on younger samples (at baseline), which may have reduced the impact of reverse causality. The majority of studies on older samples could have observed the effects of (prodromal) cognitive decline affecting physical activity rather than the other way around [25]. Similarly to Hofman et al. [62] and Rodriguez-Ayllon et al. [63], who found a bidirectional association between physical activity and brain structure, with a more consistent pattern of brain structural measures affecting physical activity, the results of our path analysis partially supported the reverse causality explanation, indicating that baseline brain health influences follow-up physical capacity, rather than baseline physical activity affecting follow-up brain health. Possible mechanisms may involve decreasing health status with age-related mitochondrial dysfunction [64, 65] and potential low-grade inflammation, which could result in fatigue [66] and a possible decline in fitness and physical capacity. Further studies are necessary to investigate this in more detail. However, our results should be interpreted with caution, due to the limited sample size, potential attenuation of effects resulting from measurement error in the assessment of physical activity/capacity and the shift from an activity-based measure at baseline to a capacity-based measure at follow-up. This change limits the interpretability of longitudinal effects, as observed associations may reflect both changes in the underlying construct being measured and true changes in the relationship over time.

### Strengths and limitations

The results of this cross-sectional observational study may be affected by various factors, including bias in self-reported physical activity [48, 49], accelerometer measurement error [67, 68] and reverse causality, among others. Moreover, our results may also be affected by the general medical status of the participants, since we did not control for other diseases within the sample. In fact, BrainAGE may reflect overall health and the cumulative impact of various factors (including previous physical activity) on brain health over an extended period of time. Furthermore, our analysis focused on only a few cognitive and structural brain measures. Although we did not observe any changes in hippocampal volume or BrainAGE, this does not rule out the possibility of changes in white matter integrity or functional connectivity. Another limitation of this observational study was the time lag between physical activity measurements and MRI scanning, which may have reduced the observed effects. Although the longitudinal design is a major strength of this study, attrition of participants at follow-up may have affected our estimates. Furthermore, the use of cross-lagged panel model design in the longitudinal setting has frequently been criticised for not distinguishing between within-person changes and between-person differences [69, 70], and our adapted design suffers from these limitations, as well as others arising from the use of different instruments to measure the construct related to physical activity at each time point (IPAQ and VSAQ). Nevertheless, compared to large volunteer-based cohorts such as the UK Biobank, the registry-based recruitment strategy of the LIFE-Adult Study may be less susceptible to healthy volunteer bias, although we cannot entirely eliminate the possibility of volunteer bias among the participants with accelerometry data in our case. Direct comparisons are limited in the absence of harmonised recruitment and assessment protocols. However, it is possible that the present study may potentially yield more generalisable estimates of the association between physical activity and brain structure and cognition for the white European population.

## Conclusion

We found no consistent positive effects of physical activity on brain structure (i.e. hippocampal volume and BrainAGE) or cognition (i.e. processing speed and executive function) in the population-based LIFE-Adult cohort. The aforementioned limitations might have reduced the strength of the potential effects. Further intervention studies with objectively measured physical activity in young and middle-aged adults are warranted to investigate the influence of physical activity on prospective brain and cognitive health. As in previous studies (e.g. [52], [53]), we observed an age-related bias in the self-reporting of physical activity. This indicates that older individuals tend to overestimate their level of activity. Future interventions targeting middle-aged adults may be necessary to raise awareness of this bias and encourage increased physical activity.

Taken together, our results are in line with those of observational longitudinal studies, which indicate minimal or no effect of physical activity on certain aspects of cognitive and/or brain health outcomes (e.g. [25, 60, 61]). Rather, they suggest a reverse pattern, whereby people with healthier brains tend to engage in more physical activities over time [62, 63].

## Data Availability

We used data from the Leipzig Research Centre for Civilization Diseases (LIFE) under project agreement PV-801. All original data will be exclusively shared by LIFE (https://www.uniklinikum-leipzig.de/einrichtungen/life) based on a project proposal in accordance with the University of Leipzig's data protection rules. The other publicly available neuroimaging datasets used in this study are accessible either openly or upon request: IXI (https://brain-development.org/ixi-dataset/), Cam-CAN (https://opendata.mrc-cbu.cam.ac.uk/projects/camcan/), Wayne State studies 10, 11, & EF datasets (https://fcon_1000.projects.nitrc.org/indi/retro/wayne_10.html, https://fcon_1000.projects.nitrc.org/indi/retro/wayne_11.html, https://fcon_1000.projects.nitrc.org/indi/retro/wayne_EF.html), the Enhanced NKI-Rockland sample (https://rocklandsample.org/for-researchers/step-3-accessing-the-data), Southwest University adult lifespan dataset (SALD; https://fcon_1000.projects.nitrc.org/indi/retro/sald.html), and Australian Imaging Biomarkers and Lifestyle study (AIBL; https://aibl.org.au/).

## Author contributions

**Polona Kalc**: Conceptualization, Formal analysis, Writing - original draft, Writing - review and editing, Visualisation. **Robert Dahnke**: Software, Data curation, Visualisation, Writing - review and editing. **Christian Sander**: Resources, Data curation, Writing - review and editing. **Frauke Beyer**: Resources, Data curation, Writing - review and editing, **Andrea Zülke**: Writing-review and editing. **Steffi G. Riedel-Heller**: Resources, Data curation, Writing - review and editing. **Veronica Witte**: Resources, Data curation, Writing – review & editing. **Christian Gaser**: Funding acquisition, Supervision, Writing - review and editing.

## Acknowledgements

We would particularly like to thank the participants and researchers involved in the LIFE-Adult study, as well as other open neuroimaging databases that were used for this manuscript (IXI, Cam-CAN, Wayne studies, Enhanced NKI-Rockland sample, and Australian Imaging Biomarkers and Lifestyle flagship study of ageing - AIBL).

We acknowledge support by the German Research Foundation Projekt-Nr. 512648189 and the Open Access Publication Fund of the Thueringer Universitaets-und Landesbibliothek Jena.

## Supplementary material

### Directed acyclic graph (DAG)

**Figure S1.**
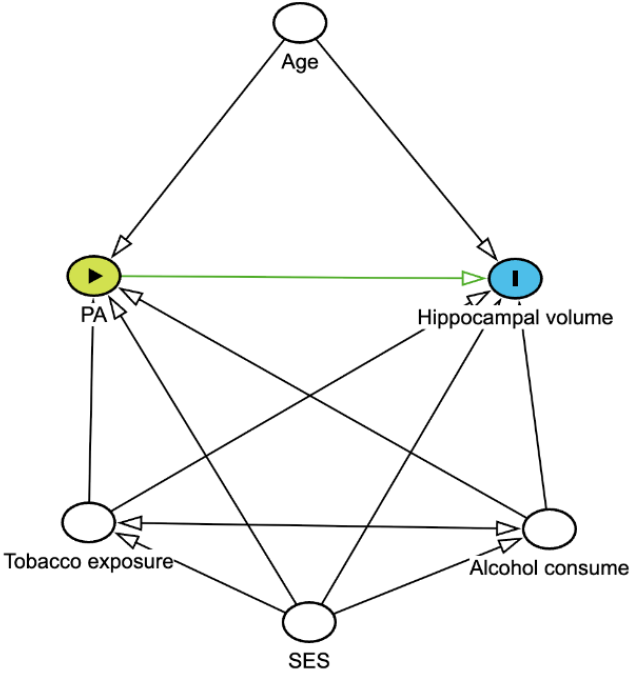
Directed acyclic graph for the effect of physical activity on hippocampal volume.

We evaluated implied independencies in the DAG employing LOESS-based conditional independence testing with bootstrap resampling (500 iterations) in the *dagitty* R package [44]. The conditional independence tests confirmed DAG-implied independencies. No statistically significant associations were observed between age and alcohol consumption (estimate: 0.028, std. error: 0.020, 95% CI [-0.009, 0.067]), SES (estimate: -0.004, std. error: 0.024, 95% CI [-0.053, 0.043]), or smoking status (estimate: -0.022, std. error: 0.022, 95% CI [-0.070, 0.019]). A minimal adjustment set for the multiple linear regression was set to age, tobacco exposure, alcohol consumption, and SES.

### BrainAGE estimation

The supervised BrainAGE machine learning model uses a Gaussian Process Regression to learn the age-related pattern from affine-registered volumetric gray matter and white matter data and outputs the predicted (brain) age. The used model was trained on a subset of scans from a large collection of healthy participants from various openly available neuroimaging datasets. The scans from the following datasets were included: IXI (https://brain-development.org/ixi-dataset/), Cam-CAN [71], Wayne State studies 10, 11, & EF datasets (Wayne State University; https://fcon_1000.projects.nitrc.org/indi/retro/wayne_10.html, https://fcon_1000.projects.nitrc.org/indi/retro/wayne_11.html, https://fcon_1000.projects.nitrc.org/indi/retro/wayne_EF.html), the Enhanced NKI-Rockland sample [72], Southwest University adult lifespan dataset (SALD; https://fcon_1000.projects.nitrc.org/indi/retro/sald.html; [73]), and Australian Imaging Biomarkers and Lifestyle study (AIBL; [74]; https://aibl.org.au/). The subset for a training sample (n = 2446, M_Age_ = 57.69 ± 14.88 years, 58% women) was chosen to match the age and sex distribution of the participants in the LIFE-Adult study. The trained algorithm was then applied to a sample of 4297 brain images from the LIFE-Adult sample, resulting in a mean absolute error (MAE) of 5.97 years and a correlation to age of 0.91. The brain age gap estimate is obtained by subtracting the chronological age from the predicted age. As the brain age gap is always overestimated for younger subjects and underestimated for older ones, an age-bias correction was applied as described in Cole [26]. Briefly, a random subset of the data was used to fit a linear regression between the chronological age as an independent and predicted age as a dependent variable. The predicted age in the LIFE-Adult dataset was then corrected by subtracting the intercept and dividing by the slope of the fitted model. This resulted in a MAE of 4.68 years. The age-bias correction is used to remove the dependency of the predicted age on age [40], we therefore did not further control for the age in the regression analyses using the BrainAGE as the outcome.

**Table S1.** Unadjusted and adjusted cross-sectional parameter estimates of self-reported total and MVPA as well as objectively measured physical activity on left hippocampal volume.

| Exposure | Outcome | Unadjusted |  |  |  |  |  |  | Adjusted |  |  |  |  |  |
| --- | --- | --- | --- | --- | --- | --- | --- | --- | --- | --- | --- | --- | --- | --- |
|  |  | n* | Estimate | SE | 95% CI | p (uncor) | p (FDR) |  | n* | Estimate | SE | 95% CI | p (uncor) | p (FDR) |
| Total PA self-report (MET hrs/day) | Left hippocampal volume | 2576-2 | -0.083 | 0.022 | -0.127, -0.039 | <.00001 | <.00001 |  | 2576-8 | -0.026<sup>a</sup> | 0.019 | -0.063, 0.012 | 0.179 | 0.269 |
| MVPA self-report (MET hrs/day) | Left hippocampal volume | 2576-2 | -0.064 | 0.022 | -0.107, -0.021 | 0.004 | 0.012 |  | 2576-8 | -0.015<sup>a</sup> | 0.019 | -0.052, 0.021 | 0.407 | 0.487 |
| Sensewear actigraphy derived average METs/day | Left hippocampal volume | 227-1 | 0.117 | 0.065 | -0.010, 0.244 | 0.071 | 0.142 |  | 227-3 | -0.046<sup>a</sup> | 0.066 | -0.175, 0.083 | 0.487 | 0.487 |

**Table S2.**
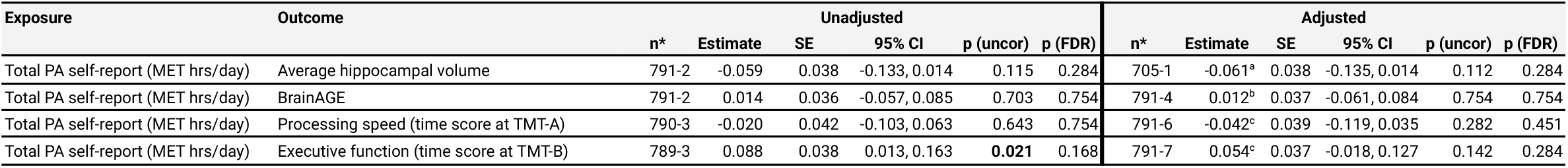
Unadjusted and adjusted cross-sectional parameter estimates of self-reported total physical activity on brain structure and cognition of adults aged 30-60 years.

| Exposure | Outcome | Unadjusted |  |  |  |  |  |  | Adjusted |  |  |  |  |  |
| --- | --- | --- | --- | --- | --- | --- | --- | --- | --- | --- | --- | --- | --- | --- |
|  |  | n* | Estimate | SE | 95% CI | p (uncor) | p (FDR) |  | n* | Estimate | SE | 95% CI | p (uncor) | p (FDR) |
| Total PA self-report (MET hrs/day) | Average hippocampal volume | 791-2 | -0.059 | 0.038 | -0.133, 0.014 | 0.115 | 0.284 |  | 705-1 | -0.061<sup>a</sup> | 0.038 | -0.135, 0.014 | 0.112 | 0.284 |
| Total PA self-report (MET hrs/day) | BrainAGE | 791-2 | 0.014 | 0.036 | -0.057, 0.085 | 0.703 | 0.754 |  | 791-4 | 0.012<sup>b</sup> | 0.037 | -0.061, 0.084 | 0.754 | 0.754 |
| Total PA self-report (MET hrs/day) | Processing speed (time score at TMT-A) | 790-3 | -0.020 | 0.042 | -0.103, 0.063 | 0.643 | 0.754 |  | 791-6 | -0.042<sup>c</sup> | 0.039 | -0.119, 0.035 | 0.282 | 0.451 |
| Total PA self-report (MET hrs/day) | Executive function (time score at TMT-B) | 789-3 | 0.088 | 0.038 | 0.013, 0.163 | 0.021 | 0.168 |  | 791-7 | 0.054<sup>c</sup> | 0.037 | -0.018, 0.127 | 0.142 | 0.284 |

**Table S3.** Unadjusted and adjusted cross-sectional parameter estimates of self-reported moderate to vigorous physical activity on brain structure and cognition in a sample aged 30-60 years.

| Exposure | Outcome | Unadjusted |  |  |  |  |  |  | Adjusted |  |  |  |  |  |
| --- | --- | --- | --- | --- | --- | --- | --- | --- | --- | --- | --- | --- | --- | --- |
|  |  | n* | Estimate | SE | 95% CI | p (uncor) | p (FDR) |  | n* | Estimate | SE | 95% CI | p (uncor) | p (FDR) |
| MVPA self-report (MET hrs/day) | Average hippocampal volume | 791-2 | -0.032 | 0.037 | -0.105, 0.042 | 0.396 | 0.671 |  | 705-1 | -0.030<sup>a</sup> | 0.038 | -0.105, 0.044 | 0.429 | 0.671 |
| MVPA self-report (MET hrs/day) | BrainAGE | 791-2 | 0.002 | 0.038 | -0.072, 0.075 | 0.968 | 0.996 |  | 791-4 | 0.000<sup>b</sup> | 0.038 | -0.074, 0.074 | 0.996 | 0.996 |
| MVPA self-report (MET hrs/day) | Processing speed (time score at TMT-A) | 790-3 | -0.027 | 0.040 | -0.105, 0.051 | 0.503 | 0.671 |  | 791-6 | -0.033<sup>c</sup> | 0.038 | -0.106, 0.041 | 0.386 | 0.671 |
| MVPA self-report (MET hrs/day) | Executive function (time score at TMT-B) | 789-3 | 0.059 | 0.037 | -0.013, 0.131 | 0.108 | 0.671 |  | 791-7 | 0.044<sup>c</sup> | 0.036 | -0.026, 0.115 | 0.219 | 0.671 |

**Table S4.**
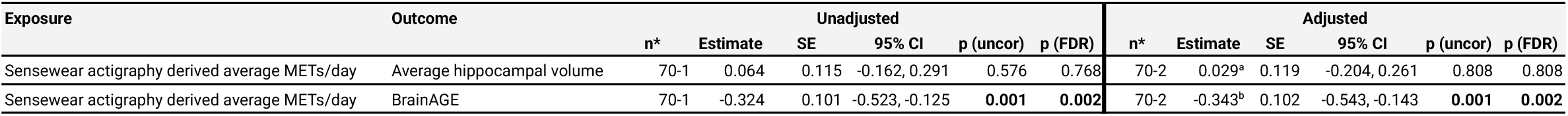
Unadjusted and adjusted cross-sectional parameter estimates of accelerometer-derived average MET/day on brain structure of adults aged 30-60 years (n = 70).

**Table S5.**
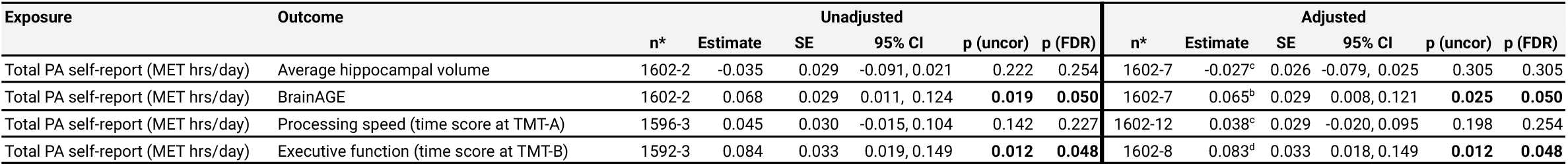
Unadjusted and adjusted cross-sectional parameter estimates of self-reported total physical activity on brain structure and cognition of adults aged >60 years.

**Table S6.** Unadjusted and adjusted cross-sectional parameter estimates of self-reported moderate to vigorous physical activity on brain structure and cognition of adults aged >60 years.

| Exposure | Outcome | Unadjusted |  |  |  |  |  | Adjusted |  |  |  |  |  |
| --- | --- | --- | --- | --- | --- | --- | --- | --- | --- | --- | --- | --- | --- |
|  |  | n* | Estimate | SE | 95% CI | p (uncor) | p (FDR) | n* | Estimate | SE | 95% CI | p (uncor) | p (FDR) |
| MVPA self-report (MET hrs/day) | Average hippocampal volume | 1602-2 | -0.036 | 0.028 | -0.091, 0.019 | 0.199 | 0.318 | 1602-7 | -0.024<sup>c</sup> | 0.026 | -0.075, 0.028 | 0.366 | 0.418 |
| MVPA self-report (MET hrs/day) | BrainAGE | 1602-2 | 0.076 | 0.029 | 0.019, 0.133 | **0.009** | 0.052 | 1602-7 | 0.072<sup>b</sup> | 0.029 | 0.015, 0.130 | **0.013** | 0.052 |
| MVPA self-report (MET hrs/day) | Processing speed (time score at TMT-A) | 1596-3 | 0.033 | 0.032 | -0.029, 0.095 | 0.293 | 0.391 | 1602-12 | 0.024<sup>c</sup> | 0.030 | -0.035, 0.083 | 0.420 | 0.420 |
| MVPA self-report (MET hrs/day) | Executive function (time score at TMT-B) | 1592-3 | 0.072 | 0.033 | 0.007, 0.137 | **0.029** | 0.077 | 1602-12 | 0.057<sup>b</sup> | 0.032 | -0.006, 0.119 | 0.076 | 0.152 |

**Table S7.** Unadjusted and adjusted cross-sectional parameter estimates of accelerometer-derived average MET/day on brain structure of older adults (age >60 years, n = 151).

| Exposure | Outcome | Unadjusted |  |  |  |  |  | Adjusted |  |  |  |  |  |
| --- | --- | --- | --- | --- | --- | --- | --- | --- | --- | --- | --- | --- | --- |
|  |  | n | Estimate | SE | 95% CI | p (uncor) | p (FDR) | n | Estimate | SE | 95% CI | p (uncor) | p (FDR) |
| Sensewear actigraphy derived average METs/day | Average hippocampal volume | 151-1 | -0.039 | 0.082 | -0.200, 0.123 | 0.638 | 0.954 | 137-1 | -0.052<sup>a</sup> | 0.081 | -0.212, 0.108 | 0.523 | 0.954 |
| Sensewear actigraphy derived average METs/day | BrainAGE | 151-1 | 0.005 | 0.079 | -0.149, 0.158 | 0.954 | 0.954 | 151-3 | -0.006<sup>b</sup> | 0.076 | -0.155, 0.144 | 0.942 | 0.954 |

**Table S8.** Unadjusted and adjusted cross-sectional parameter estimates of self-reported total physical activity on brain structure and cognition of **men**.

| Exposure | Outcome | Unadjusted |  |  |  |  |  | Adjusted |  |  |  |  |  |
| --- | --- | --- | --- | --- | --- | --- | --- | --- | --- | --- | --- | --- | --- |
|  |  | n* | Estimate | SE | 95% CI | p (uncor) | p (FDR) | n* | Estimate | SE | 95% CI | p (uncor) | p (FDR) |
| Total PA self-report (MET hrs/day) | Average hippocampal volume | 1370-2 | -0.099 | 0.003 | -0.158, -0.040 | **0.001** | **0.004** | 1370-7 | -0.047<sup>a</sup> | 0.024 | -0.094, 0.000 | **0.049** | 0.082 |
| Total PA self-report (MET hrs/day) | BrainAGE | 1370-2 | 0.056 | 0.029 | -0.002, 0.113 | 0.057 | 0.082 | 1370-7 | 0.054<sup>b</sup> | 0.030 | -0.003, 0.112 | 0.065 | 0.082 |
| Total PA self-report (MET hrs/day) | Processing speed (time score at TMT-A) | 1363-3 | 0.061 | 0.034 | -0.005, 0.128 | 0.072 | 0.082 | 1370-12 | 0.003<sup>c</sup> | 0.029 | -0.053, 0.060 | 0.914 | 0.914 |
| Total PA self-report (MET hrs/day) | Executive function (time score at TMT-B) | 1360-3 | 0.125 | 0.034 | 0.059, 0.191 | **<.00001** | **<.00001** | 1370-12 | 0.055<sup>c</sup> | 0.029 | -0.002, 0.113 | 0.059 | 0.082 |

**Table S9.** Unadjusted and adjusted cross-sectional parameter estimates of self-reported total physical activity on brain structure and cognition of **women**.

| Exposure | Outcome | Unadjusted |  |  |  |  |  | Adjusted |  |  |  |  |  |
| --- | --- | --- | --- | --- | --- | --- | --- | --- | --- | --- | --- | --- | --- |
|  |  | n* | Estimate | SE | 95% CI | p (uncor) | p (FDR) | n* | Estimate | SE | 95% CI | p (uncor) | p (FDR) |
| Total PA self-report (MET hrs/day) | Average hippocampal volume | 1202-2 | -0.062 | 0.303 | -0.128, 0.003 | **0.061** | 0.163 | 1202-6 | -0.005<sup>a</sup> | 0.030 | -0.064, 0.054 | 0.865 | 0.865 |
| Total PA self-report (MET hrs/day) | BrainAGE | 1202-2 | 0.027 | 0.032 | -0.036, 0.090 | 0.395 | 0.604 | 1202-6 | 0.018<sup>b</sup> | 0.032 | -0.045, 0.082 | 0.572 | 0.654 |
| Total PA self-report (MET hrs/day) | Processing speed (time score at TMT-A) | 1202-3 | 0.092 | 0.034 | 0.026, 0.159 | **0.007** | **0.028** | 1202-8 | 0.022<sup>c</sup> | 0.030 | -0.036, 0.080 | 0.453 | 0.604 |
| Total PA self-report (MET hrs/day) | Executive function (time score at TMT-B) | 1200-3 | 0.116 | 0.039 | 0.039, 0.193 | **0.003** | **0.024** | 1202-9 | 0.027<sup>c</sup> | 0.036 | -0.042, 0.097 | 0.442 | 0.604 |

**Table S10.** Unadjusted and adjusted cross-sectional parameter estimates of accelerometer-derived average MET/day on brain structure of **men**.

| Exposure | Outcome | Unadjusted |  |  |  |  |  | Adjusted |  |  |  |  |  |
| --- | --- | --- | --- | --- | --- | --- | --- | --- | --- | --- | --- | --- | --- |
|  |  | n* | Estimate | SE | 95% CI | p (uncor) | p (FDR) | n* | Estimate | SE | 95% CI | p (uncor) | p (FDR) |
| Senswear actigraphy derived average METs/day | Average hippocampal volume | 117-1 | 0.120 | 0.080 | -0.037, 0.277 | 0.134 | 0.536 | 117-2 | -0.072<sup>a</sup> | 0.080 | -0.228, 0.085 | 0.368 | 0.736 |
| Senswear actigraphy derived average METs/day | BrainAGE | 117-1 | 0.042 | 0.097 | -0.148, 0.232 | 0.667 | 0.760 | 117-2 | 0.030<sup>b</sup> | 0.098 | -0.162, 0.222 | 0.760 | 0.760 |

**Table S11.** Unadjusted and adjusted cross-sectional parameter estimates of accelerometer-derived average MET/day on brain structure of **women**.

| Exposure | Outcome | Unadjusted |  |  |  |  |  | Adjusted |  |  |  |  |  |
| --- | --- | --- | --- | --- | --- | --- | --- | --- | --- | --- | --- | --- | --- |
|  |  | n* | Estimate | SE | 95% CI | p (uncor) | p (FDR) | n* | Estimate | SE | 95% CI | p (uncor) | p (FDR) |
| Senswear actigraphy derived average METs/day | Average hippocampal volume | 110-1 | 0.171 | 0.099 | -0.022, 0.365 | 0.083 | 0.115 | 110-3 | 0.071<sup>a</sup> | 0.097 | -0.119, 0.262 | 0.462 | 0.462 |
| Senswear actigraphy derived average METs/day | BrainAGE | 110-1 | -0.154 | 0.087 | -0.325, 0.017 | 0.078 | 0.115 | 110-3 | -0.148<sup>b</sup> | 0.086 | -0.316, 0.021 | 0.086 | 0.115 |

**Table S12.**
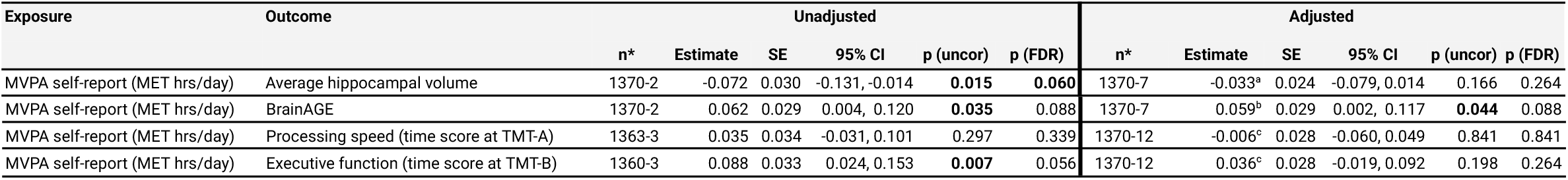
Unadjusted and adjusted cross-sectional parameter estimates of self-reported moderate to vigorous physical activity on brain structure and cognition in **men**.

**Table S13.** Unadjusted and adjusted cross-sectional parameter estimates of self-reported moderate to vigorous physical activity on brain structure and cognition in **women**.

| Exposure | Outcome | Unadjusted |  |  |  |  |  | Adjusted |  |  |  |  |  |
| --- | --- | --- | --- | --- | --- | --- | --- | --- | --- | --- | --- | --- | --- |
|  |  | n* | Estimate | SE | 95% CI | p (uncor) | p (FDR) | n* | Estimate | SE | 95% CI | p (uncor) | p (FDR) |
| MVPA self-report (MET hrs/day) | Average hippocampal volume | 1202-2 | -0.052 | 0.033 | -0.116, 0.012 | 0.111 | 0.296 | 1202-6 | -0.005<sup>a</sup> | 0.029 | -0.063, 0.052 | 0.857 | 0.857 |
| MVPA self-report (MET hrs/day) | BrainAGE | 1202-2 | 0.027 | 0.033 | -0.038, 0.091 | 0.216 | 0.432 | 1202-6 | 0.019<sup>b</sup> | 0.033 | -0.045, 0.083 | 0.559 | 0.646 |
| MVPA self-report (MET hrs/day) | Processing speed (time score at TMT-A) | 1202-3 | 0.071 | 0.037 | -0.002, 0.144 | 0.056 | 0.224 | 1202-8 | 0.019<sup>c</sup> | 0.032 | -0.045, 0.082 | 0.565 | 0.646 |
| MVPA self-report (MET hrs/day) | Executive function (time score at TMT-B) | 1200-3 | 0.096 | 0.041 | 0.017, 0.176 | **0.017** | 0.136 | 1202-9 | 0.031<sup>c</sup> | 0.037 | -0.041, 0.103 | 0.397 | 0.635 |

\* Number of subjects - missing data patterns
<sup>a</sup> Adjusted for linear and quadratic effect of age, tobacco exposure, alcohol consumption, socioeconomic status
<sup>b</sup> Adjusted for tobacco exposure, alcohol consumption, socioeconomic status
<sup>c</sup> Adjusted for linear effect of age, tobacco exposure, alcohol consumption, socioeconomic status
<sup>d</sup> Adjusted for tobacco exposure and alcohol consumption

